# Influence of vitamin D supplementation on muscle strength and exercise capacity in Mongolian schoolchildren: a randomised controlled trial

**DOI:** 10.1101/2024.03.27.24304943

**Authors:** Davaasambuu Ganmaa, Stephanie Hemmings, David A. Jolliffe, Uyanga Buyanjargal, Gantsetseg Garmaa, Unaganshagai Adiya, Tumenulzii Tumurbaatar, Khulan Dorjnamjil, Enkhtsetseg Tserenkhuu, Sumiya Erdenenbaatar, Enkhjargal Tsendjav, Nomin Enkhamgalan, Chuluun-Erdene Achtai, Yagaantsetseg Talhaasuren, Tuya Byambasuren, Erdenetuya Ganbaatar, Erkhembulgan Purevdorj, Adrian R Martineau

## Abstract

**Objective:** To determine whether weekly oral vitamin D supplementation influences grip strength, explosive leg power, cardiorespiratory fitness or spirometric lung volumes in Mongolian schoolchildren.

**Methods:** Multicentre, randomised, double-blind, placebo-controlled clinical trial conducted in children aged 6-13 years at baseline attending 18 schools in Ulaanbaatar. The intervention was weekly oral doses of 14,000 IU vitamin D_3_ (n=4418) or placebo (n=4433) for 3 years. Outcome measures were grip strength, standing long jump distance and serum 25-hydroxyvitamin D (25[OH]D) concentrations (determined in all participants), peak oxygen uptake (VO_2peak_, determined in a subset of 632 participants using 20-metre multi-stage shuttle run tests) and spirometric outcomes (determined in a subset of 1,343 participants).

**Results:** 99.8% of participants had serum 25(OH)D concentrations <75 nmol/L at baseline, and mean end-study 25(OH)D concentrations in children randomised to vitamin D vs. placebo were 77.4 vs. 26.7 nmol/L (mean difference 50.7 nmol/L, 95% CI, 49.7 to 51.4). However, vitamin D supplementation did not influence mean grip strength, standing long jump distance, VO_2peak_, spirometric lung volumes or peak expiratory flow rate, either overall or within sub-groups defined by sex, baseline 25(OH)D concentration <25 vs. ≥25 nmol/L or calcium intake <500 vs. ≥500 mg/day.

**Conclusion:** A 3-year course of weekly oral supplementation with 14,000 IU vitamin D_3_ elevated serum 25(OH)D concentrations in Mongolian schoolchildren with a high baseline prevalence of vitamin D deficiency. However, this intervention did not influence grip strength, explosive leg power, peak oxygen uptake or spirometric lung volumes, either overall or in sub-group analyses.

**KEY MESSAGES:** **What is already known on this topic?**

Observational studies have reported that vitamin D deficiency associates with reduced muscle strength and peak oxygen uptake in children, but randomised controlled trials (RCT) of vitamin D supplementation to improve grip strength and cardiorespiratory fitness in this age-group have yielded conflicting results.

**What this study adds**

This Phase 3 multicentre RCT of vitamin D supplementation, conducted in Mongolian schoolchildren with a high baseline prevalence of asymptomatic vitamin D deficiency, found that a 3-year course of weekly oral supplementation with 14,000 IU vitamin D_3_ was effective in elevating serum 25-hydroxyvitamin D concentrations. However this intervention did not influence participants’ grip strength, long jump distance, peak oxygen uptake, spirometric lung volumes or peak expiratory flow rate, either overall or in sub-group analyses.

**How this study might affect research, practice or policy**

Taken together with results from another Phase 3 randomised controlled trial of vitamin D supplementation conducted in South African children, our findings do not suggest a role for weekly oral vitamin D supplementation to enhance muscle strength, peak oxygen uptake or respiratory function in schoolchildren in whom rickets has been excluded.

## INTRODUCTION

Muscular strength and exercise tolerance in childhood are positive correlates of physical and mental health that associate with reduced cardiometabolic risk later in life.^1–4^ Vitamin D plays a key role in supporting normal development and function of skeletal muscle, the lung and the heart.^5–7^ Vitamin D deficiency is common among children living in higher- and lower-income countries alike,^8–11^ and has been reported to associate with lower muscle strength, poorer cardiorespiratory fitness and lower spirometric lung volumes in children and adolescents participating in observational studies.^12–14^ Numerous randomised controlled trials (RCTs) of vitamin D supplementation to improve muscle strength and power have been conducted in adults, with meta-analysis revealing small positive impacts.^15^ Fewer RCTs have been conducted in children, and these have yielded inconsistent results. One trial in male soccer players in Tunisia aged 8-15 years has reported improvements in jump, sprint and shuttle run outcomes,^16^ while another conducted in 12-14 year-old girls in the United Kingdom reported a statistically significant improvement in efficiency of movement, with trends towards improvements in jumping velocity and grip strength.^17^ Other RCTs conducted in children and adolescents living in Denmark,^18^ ^19^ the United States,^20^ Israel ^21^ and Lebanon^22^ have reported null overall effects for outcomes including grip strength, leg press strength and swimming performance. No such trials have yet been conducted in Asia; moreover, there is a lack of large, multicentre trials examining the impact of prolonged (greater than one year) vitamin D supplementation on muscle strength, cardiorespiratory fitness and spirometric outcomes in children with a high baseline prevalence of vitamin D deficiency, regardless of setting.

An opportunity to address this gap in the literature arose during conduct of a phase 3 randomised controlled trial of vitamin D supplementation in 8,851 school children aged 6 – 13 years living in Mongolia who were given weekly oral vitamin D supplementation over 3 years. ^23–25^ The primary aim of the trial was to test whether this intervention reduced risk of incident tuberculosis infection; null results for this outcome have been reported elsewhere.^23^ This paper reports findings for secondary outcomes including grip strength and standing long jump distance, measured at annual intervals in all participants; peak oxygen uptake (VO_2peak_), estimated using 20 metre shuttle run tests performed at annual intervals in a subset of 632 participants; and spirometric outcomes, measured at 3-year follow-up in a subset of 1,343 participants.

## METHODS

### Study Design, Setting, Participants and Randomisation

We conducted a parallel two-arm double-blind individually randomised placebo-controlled trial in eighteen public schools in Ulaanbaatar, Mongolia, as previously described.^23–25^ The primary outcome of the trial was acquisition of latent tuberculosis infection; the current manuscript reports effects of the intervention on pre-specified secondary outcomes relating to grip strength, explosive leg power, peak oxygen uptake and spirometric outcomes. Principal inclusion criteria were age 6-13 years at screening and attendance at a participating school; principal exclusion criteria were a positive QuantiFERON-TB Gold in-tube assay (QFT) result, presence of conditions associated with vitamin D hypersensitivity (primary hyperparathyroidism or sarcoidosis) or immunocompromise (taking immunosuppressant medication or cytotoxic therapy), use of vitamin D supplements, signs of rickets (all participants were screened for rickets via physical examination by a paediatrician), or intention to move from Ulaanbaatar within 4 years of enrolment. Eligible participants were individually randomised to receive a weekly capsule containing 14,000 IU (350 μg) vitamin D_3_ or placebo for three years, with a one-to-one allocation ratio and stratification by school of attendance. Treatment allocation was concealed from participants, care providers, and all trial staff to maintain the double-blind. The trial is registered with clinicalTrials.gov (NCT02276755).

### Baseline Procedures

At baseline, participants’ parents were asked to complete questionnaires detailing their socioeconomic circumstances, lifestyle and dietary factors influencing vitamin D status, and intake of foods previously shown to be major contributors to dietary calcium intake in urban Mongolia.^26^ For all participants, height was then measured using a portable stadiometer (SECA, Hamburg, Germany), weight was measured using a Digital Floor Scale (SECA), grip strength was measured as described elsewhere^27^ using a portable dynamometer (Takei Digital Grip Strength Dynamometer, Model T.K.K.5401, with the best of 2 readings for the dominant hand recorded, except where injury precluded measurement, where strength of the other hand was measured), and standing long jump distance was measured as described elsewhere^28^ using a DiCUNO measuring tape, with the best of 2 readings recorded. In a subset of 620 children who additionally participated in the exercise sub-study, a 20-metre multi-stage shuttle run test was administered as described below to determine VO2peak. A blood sample was drawn from all participants at baseline for separation and storage of serum for determination of baseline 25(OH)D concentrations as described below.

### Follow-Up Procedures and Outcomes

During school terms, study participants had weekly face-to-face visits at which study capsules were administered and adverse events were recorded. During school holidays, children were either given a single bolus dose of up to 36,000 IU (shorter holidays), study staff travelled to participants homes to administer medication, or parents were supplied with sufficient trial medication to cover the holiday period, along with instructions on its storage and administration. Using the same methods as at baseline, all participants were re-assessed at 12-, 24- and 36-month follow-up for grip strength and long jump distance. Exercise sub-study participants additionally completed 20-metre multi-stage shuttle run tests at 12-, 24- and 36-month follow-up, as described below. Spirometry sub-study participants additionally underwent spirometry testing at 36-month follow-up using a portable spirometer (spirolab III, Medical International Research, Rome, Italy) and performed according to ERS/ATS standards^29^ to assess % predicted forced expiratory volume in one second (FEV1), % predicted forced vital capacity (FVC), % predicted FEV1/FVC, % predicted peak expiratory flow rate (PEFR) and % predicted forced expiratory flow over the middle one-half of the FVC (FEF25-75).

### Measurement of vitamin D status

25(OH)D concentrations were determined in serum samples from baseline and 3-year follow-up using an enzyme-linked fluorescent assay (VIDAS 25OH Vitamin D total, bioMérieux, Marcy-l’Étoile, France). Non-zero 25(OH)D values were standardised using a published method,^30^ utilising a set of 40 serum samples provided by DEQAS (the Vitamin D External Quality Assessment Scheme, http://www.deqas.org/). Total coefficient of variation (CV) was 7.9%, mean bias was 7.7% and the limit of quantitation (LOQ) was 20.2 nmol/L. Values below the LOQ were classified as < 20.2 nmol/L.

### Exercise test

A 20-metre multi-stage shuttle run test was conducted using freely available recorded instruction (Shuttle run bleep test, www.bleeptests.com). Two lines were marked 20 meters apart and an audible ‘beep’ signalled to participants the speed required to run between them. Participants’ number of completed laps were used to derive their estimated VO_2peak_ using a published formula.^31^

### Sample Size and Statistical Methods

The sample size calculation for the main trial was based on the power needed to detect a clinically significant effect of the intervention on the primary endpoint (incident latent tuberculosis infection) as described previously [20]. For the exercise sub-study, assuming a standard deviation for VO_2peak_ of 6.5 ml/kg/min at 3-year follow-up,^31^ we calculated that a total of 334 participants (167 per arm) would need to be recruited and followed up to have 80% power to detect a clinically significant difference of 2 ml/kg/min between arms with 5% alpha. Allowing for 20% loss to follow-up, we originally estimated that a total of 420 participants (210 per arm) would need to be recruited to the exercise sub-study. Subsequently, we became concerned that rates of loss to follow-up might be higher than originally anticipated, and consequently the target sample size for this sub-study was increased to 614. Ultimately, the exercise sub-study over-recruited slightly (n=632). Spirometry was performed for a subset of children who participated in a bone health sub-study, powered to detect an effect of the intervention on radial speed of sound Z-scores as described elsewhere.^25^

Estimated calcium intakes were calculated on the basis of parental responses to a food frequency questionnaire, as described elsewhere.^24^. Anthropometric measurements and data on participants’ age and sex were used to compute Z-scores for height-for-age and BMI-for-age, using the Canadian Pediatric Endocrine Group who2007 Shiny App (https://cpeg-gcep.shinyapps.io/who2007_cpeg/) based on WHO 2007 growth reference data for 5-19 years.^32^ Serum 25(OH)D values were adjusted for seasonal variation prior to analysis using a sinusoidal model.^33^ Outcomes measured over multiple years’ follow-up were analyzed overall and in each sub-group using mixed models for repeated measures with fixed effects for treatment and time and treatment-by-time interaction, adjusted for school of attendance and random effects for individuals. Adjusted treatment mean differences at different time-points are presented with 95% confidence intervals, and significance tests were conducted for the treatment effect at each time point and the overall treatment-by-time interaction. Where overall P values were less than 0.05, we applied a Benjamini Hochberg procedure with a 10% false discovery rate ^34^ to the relevant family of P values to adjust for multiple comparisons. Outcomes measured at end-study only were analyzed using general linear models, adjusted for school of attendance. Pre-specified sub-group analyses were conducted according to participants’ sex (males vs. females), baseline deseasonalised 25(OH)D concentration (<25 nmol/L vs. ≥25 nmol/L) and estimated calcium intake (<500 mg/day vs. ≥500 mg/day). The primary P-values for outcome modelling were the overall P values, i.e. those associated with the interaction between follow-up timepoint and treatment allocation.

### Patient and Public Involvement

We consulted children and their parents / guardians on questionnaire design and acceptability of clinical measurements prior to implementation of the trial. Study findings were disseminated via community engagement events.

## RESULTS

### Participants

11,475 children were invited to participate in the study from September 2015 to March 2017 inclusive, of whom 9,814 underwent QFT testing: 8,851 QFT-negative children were randomly assigned to receive vitamin D or placebo (4418 vs. 4433, respectively) as previously described.^23^ Of these, 632 children (302 vs. 330 randomised to vitamin D vs. placebo, respectively) also participated in the exercise sub-study, and 1343 children (666 vs. 677 randomised to vitamin D vs. placebo, respetively) underwent spirometry at 3-year follow-up (Figure 1). Table 1 shows participants’ baseline characteristics: overall, mean age was 9.4 years and 49.3% were female. Mean deseasonalised baseline serum 25(OH)D concentration was 29.7 nmol/L (SD 10.5). Baseline characteristics were well balanced between participants randomised to vitamin D vs. placebo, both for those in the main trial and for participants contributing data to the exercise and spirometry sub-studies. Mean serum 25(OH)D concentration at 3-year follow-up was higher in the vitamin D group vs. the placebo group (77.4 vs. 26.7 nmol/L, respectively; mean difference 50.7 nmol/L, 95% CI, 49.7 to 51.4). 8549 participants contributed data to the analysis of grip strength; 8539 to the analysis of long jump distance; 611 to the analysis of cardiorespiratory fitness; and 1343 to the analysis of spirometric outcomes (Figure 1).

**Figure 1:**
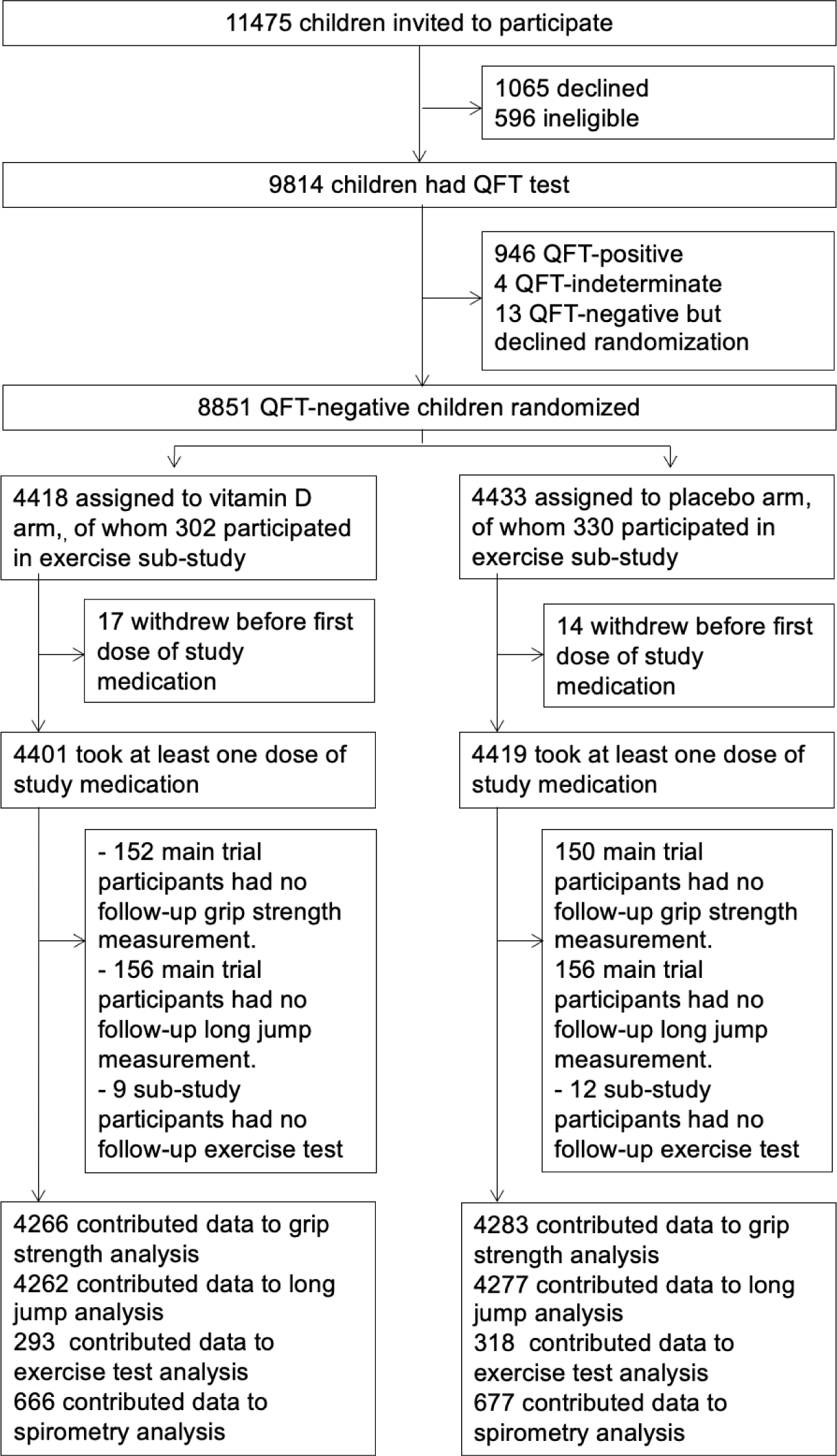
Trial profile.

**Table 1:**
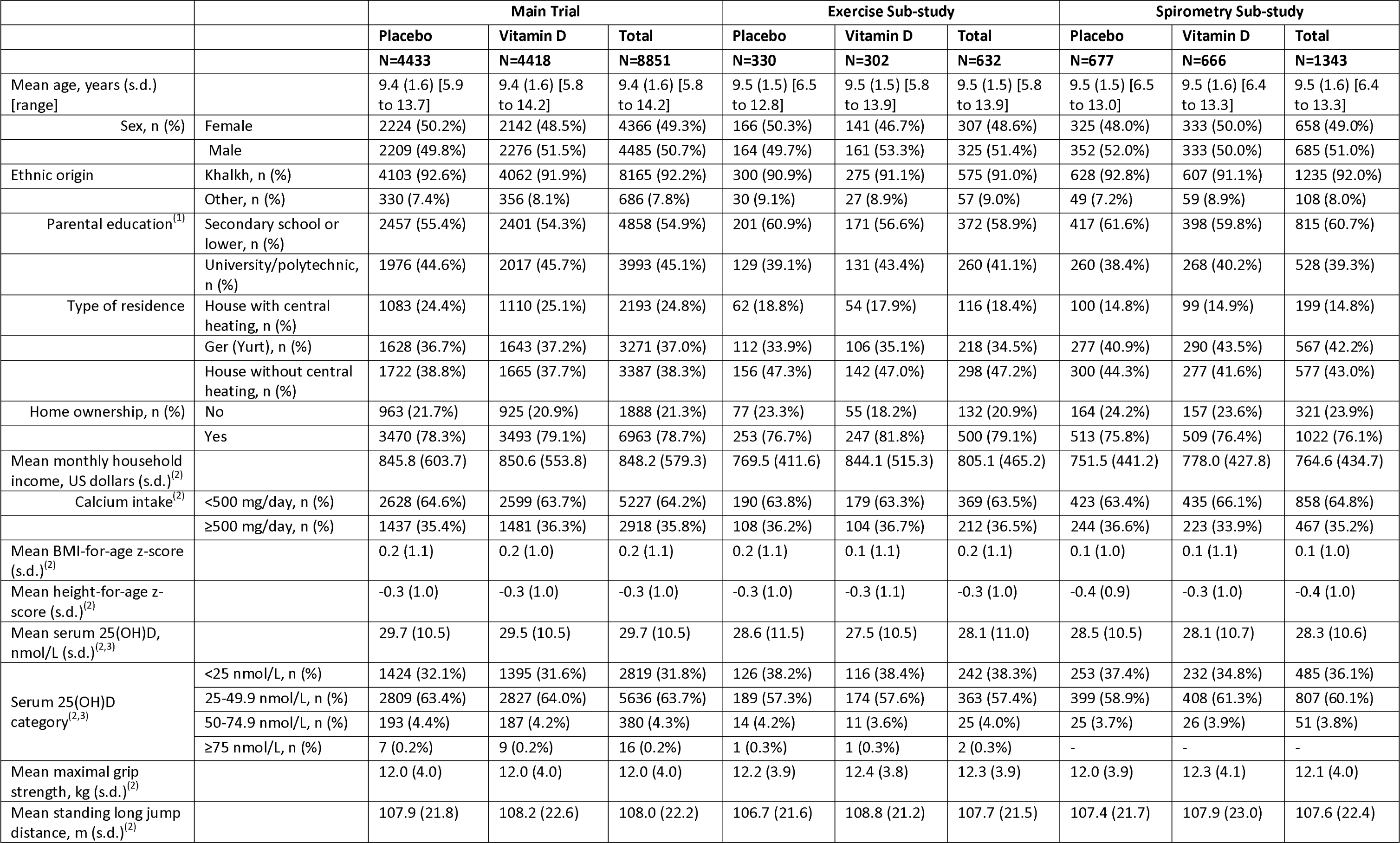
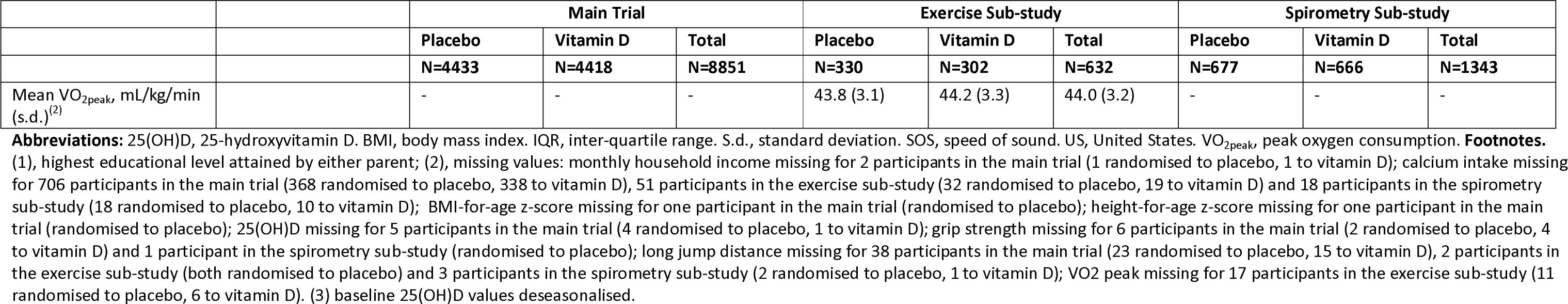
Participants’ baseline characteristics by allocation and overall: main trial, exercise sub-study and spirometry sub-study.

### Outcomes

Allocation to vitamin D vs. placebo did not influence mean grip strength, either overall or in sub-groups defined by male vs. female sex, baseline 25(OH)D concentration <25 vs. ≥25 nmol/L or estimated calcium intake <500 vs ≥500 mg/day (Table 2). Similarly, no effect of the intervention was seen on long jump distance, either overall or by sub-group, after correction for multiple comparisons testing (Table 3). Among exercise sub-study participants, allocation to vitamin D vs. placebo did not influence mean VO_2peak_, either overall or within sub-groups defined by male vs. female sex, baseline 25(OH)D concentration <25 vs. ≥25 nmol/L or estimated calcium intake <500 vs ≥500 mg/day (Table 4). Among participants who underwent spirometry at 3-year follow-up, allocation to vitamin D vs. placebo did not influence % predicted FEV1, FVC, FEV1/FVC, PEFR or FEF25-75 after correction for multiple comparisons testing, either overall or within sub-groups defined by male vs. female sex, baseline 25(OH)D concentration <25 vs. ≥25 nmol/L or estimated calcium intake <500 vs ≥500 mg/day (Table 5).

**Table 2.** Mean grip strength in main trial participants at 1-, 2- and 3-year follow-up by allocation: overall and by sub-group.

|  |  |  | Vitamin D arm:<br>mean strength,<br>kg (s.d.) [n] | Placebo arm:<br>mean strength,<br>kg (s.d.) [n] | Adjusted mean<br>difference (95% CI)<br><sup>(1)</sup> | P for<br>timepoint | Overall<br>P | P for<br>interaction |
| --- | --- | --- | --- | --- | --- | --- | --- | --- |
| Overall |  | 1<br>year | 15.0 (4.9)<br>[4,219] | 14.9 (4.8)<br>[4,229] | 0.03 (-0.19, 0.25) | 0.79 | 0.999 | -- |
|  |  | 2<br>years | 17.1 (5.5)<br>[3,802] | 17.3 (5.5)<br>[3,829] | -0.09 (-0.31, 0.13) | 0.41 |  |  |
|  |  | 3<br>years | 20.0 (6.4)<br>[4,074] | 20.0 (6.4)<br>[4,054] | 0.01 (-0.21, 0.23) | 0.94 |  |  |
| By sex | Male | 1<br>year | 15.6 (5.0)<br>[2,184] | 15.5 (4.8)<br>[2,103] | 0.10 (-0.22, 0.42) | 0.53 | 0.90 | 0.55 |
|  |  | 2<br>years | 17.9 (5.8)<br>[1,952] | 18.0 (5.8)<br>[1,902] | -0.07 (-0.40, 0.26) | 0.67 |  |  |
|  |  | 3<br>years | 21.4 (7.2)<br>[2,100] | 21.4 (7.1)<br>[2,010] | -0.00 (-0.32, 0.32) | 0.99 |  |  |
|  | Female | 1<br>year | 14.2 (4.7)<br>[2,035] | 14.3 (4.7)<br>[2,126] | -0.10 (-0.38, 0.18) | 0.47 | 0.48 |  |
|  |  | 2<br>years | 16.2 (5.1)<br>[1,850] | 16.5 (5.1)<br>[1,927] | -0.19 (-0.47, 0.09) | 0.19 |  |  |
|  |  | 3<br>years | 18.6 (5.1)<br>[1,974] | 18.7 (5.2)<br>[2,044] | -0.10 (-0.38, 0.18) | 0.5 |  |  |
| By baseline<br>25(OH)D<br>concentration<sup>(2)</sup> | <25<br>nmol/L | 1<br>year | 15.5 (5.2)<br>[1,342] | 15.5 (5.3)<br>[1,352] | 0.05 (-0.33, 0.44) | 0.78 | 0.73 | 0.75 |
|  |  | 2<br>years | 17.7 (5.9)<br>[1,214] | 17.9 (6.0)<br>[1,226] | -0.05 (-0.44, 0.34) | 0.81 |  |  |
|  |  | 3<br>years | 20.4 (6.7)<br>[1,291] | 20.5 (6.7)<br>[1,309] | 0.14 (-0.24, 0.53) | 0.47 |  |  |
|  | ≥25<br>nmol/L | 1<br>year | 14.7 (4.7)<br>[2,876] | 14.6 (4.5)<br>[2,873] | 0.08 (-0.17, 0.33) | 0.53 | 0.77 |  |
|  |  | 2<br>years | 16.8 (5.3)<br>[2,587] | 16.9 (5.3)<br>[2,600] | -0.05 (-0.31, 0.21) | 0.71 |  |  |
|  |  | 3<br>years | 19.8 (6.3)<br>[2,782] | 19.8 (6.2)<br>[2,742] | 0.02 (-0.24, 0.27) | 0.91 |  |  |
| By calcium intake | <500<br>mg/day | 1<br>year | 15.0 (4.8)<br>[1,463] | 14.9 (4.8)<br>[1,422] | 0.12 (-0.16, 0.40) | 0.41 | 0.51 | 0.20 |
|  |  | 2<br>years | 17.2 (5.6)<br>[1,355] | 17.2 (5.6)<br>[1,305] | -0.00 (-0.29, 0.29) | 0.998 |  |  |
|  |  | 3<br>years | 20.2 (6.4)<br>[1,444] | 20.0 (6.5)<br>[1,410] | 0.18 (-0.10, 0.46) | 0.21 |  |  |
|  | ≥500<br>mg/day | 1<br>year | 14.8 (5.0)<br>[2,572] | 15.0 (4.7)<br>[2,596] | 0.12 (-0.16, 0.40) | 0.41 | 0.38 |  |
|  |  | 2<br>years | 16.8 (5.5)<br>[2,336] | 17.3 (5.5)<br>[2,400] | -0.00 (-0.29, 0.29) | 0.998 |  |  |
|  |  | 3<br>years | 19.7 (6.5)<br>[2,549] | 20.1 (6.2)<br>[2,561] | 0.18 (-0.10, 0.46) | 0.21 |  |  |

**Abbreviations:** 25(OH)D, 25-hydroxyvitamin D. CI, confidence interval. S.d., standard deviation. N, number.
**Footnotes.** (1) adjusted for baseline value and school of attendance. (2) deseasonalised values.

**Table 3.** Mean long jump distance in main trial participants at 1-, 2- and 3-year follow-up by allocation: overall and by sub-group.

|  |  |  | Vitamin D arm:<br>mean distance,<br>m (s.d.) [n] | Placebo arm:<br>mean distance,<br>m (s.d.) [n] | Adjusted mean<br>difference (95% CI)<br><sup>(1)</sup> | P for<br>timepoint | Overall<br>P | P for<br>interaction |
| --- | --- | --- | --- | --- | --- | --- | --- | --- |
| Overall |  | 1<br>year | 117.6 (22.4)<br>[4,206] | 117.1 (21.9)<br>[4,201] | 0.47 (-0.50, 1.45) | 0.34 | 0.33 | -- |
|  |  | 2<br>years | 124.2 (23.5)<br>[3,790] | 124.0 (22.8)<br>[3,809] | 0.42 (-0.57, 1.41) | 0.41 |  |  |
|  |  | 3<br>years | 132.0 (26.4)<br>[4,012] | 131.3 (25.4)<br>[3,988] | 0.76 (-0.22, 1.74) | 0.13 |  |  |
| By sex | Male | 1<br>year | 125.9 (22.2)<br>[2,177] | 124.9 (21.6)<br>[2,094] | 0.71 (-0.66, 2.08) | 0.31 | 0.17 | 0.04<sup>(2)</sup> |
|  |  | 2<br>years | 134.2 (23.7)<br>[1,944] | 133.1 (23.0)<br>[1,895] | 1.10 (-0.30, 2.50) | 0.13 |  |  |
|  |  | 3<br>years | 144.4 (26.9)<br>[2,065] | 142.9 (25.8)<br>[1,984] | 1.32 (-0.06, 2.70) | 0.06 |  |  |
|  | Female | 1<br>year | 108.8 (18.9)<br>[2,029] | 109.4 (19.2)<br>[2,107] | -0.40 (-1.53, 0.72) | 0.48 | 0.18 |  |
|  |  | 2<br>years | 113.6 (18.0)<br>[1,846] | 115.0 (18.6)<br>[1,914] | -1.01 (-2.17, 0.14) | 0.09 |  |  |
|  |  | 3<br>years | 118.8 (18.3)<br>[1,947] | 119.9 (18.9)<br>[2,004] | -0.73 (-1.87, 0.41) | 0.21 |  |  |
| By baseline<br>25(OH)D<br>concentration<sup>(3)</sup> | <25<br>nmol/L | 1<br>year | 119.2 (22.6)<br>[1,339] | 119.3 (22.1)<br>[1,343] | 0.32 (-1.42, 2.05) | 0.72 | 0.78 | 0.54 |
|  |  | 2<br>years | 125.6 (24.1)<br>[1,212] | 126.1 (23.9)<br>[1,219] | 0.51 (-1.26, 2.28) | 0.58 |  |  |
|  |  | 3<br>years | 133.3 (26.8)<br>[1,268] | 133.3 (25.4)<br>[1,288] | 0.80 (-0.96, 2.55) | 0.37 |  |  |
|  | ≥25<br>nmol/L | 1<br>year | 116.9 (22.2)<br>[2,866] | 116.1 (21.7)<br>[2,854] | 0.65 (-0.51, 1.81) | 0.27 | 0.23 |  |
|  |  | 2<br>years | 123.5 (23.2)<br>[2,577] | 123.0 (22.2)<br>[2,587] | 0.48 (-0.71, 1.67) | 0.43 |  |  |
|  |  | 3<br>years | 131.4 (26.2)<br>[2,743] | 130.4 (25.3)<br>[2,697] | 0.85 (-0.33, 2.02) | 0.16 |  |  |
| By calcium intake | <500<br>mg/day | 1<br>year | 117.7 (22.0)<br>[1,461] | 117.3 (21.9)<br>[1,412] | 0.34 (-0.90, 1.59) | 0.59 | 0.42 | 0.84 |
|  |  | 2<br>years | 124.3 (23.3)<br>[1,355] | 123.8 (22.6)<br>[1,301] | 0.55 (-0.72, 1.81) | 0.40 |  |  |
|  |  | 3<br>years | 131.8 (26.2)<br>[1,422] | 131.1 (25.2)<br>[1,382] | 0.65 (-0.60, 1.89) | 0.31 |  |  |
|  | ≥500<br>mg/day | 1<br>year | 117.5 (22.8)<br>[2,562] | 116.7 (21.9)<br>[2,579] | 0.96 (-0.76, 2.67) | 0.28 | 0.49 |  |
|  |  | 2<br>years | 124.0 (23.9)<br>[2,329] | 124.3 (23.1)<br>[2,385] | 0.31 (-1.44, 2.05) | 0.73 |  |  |
|  |  | 3<br>years | 132.5 (27.0)<br>[2,510] | 131.7 (25.7)<br>[2,527] | 1.30 (-0.42, 3.03) | 0.14 |  |  |

**Abbreviations:** 25(OH)D, 25-hydroxyvitamin D. CI, confidence interval. S.d., standard deviation. N, number.
**Footnotes.** (1) adjusted for baseline value and school of attendance. (2) after correction for multiple comparisons testing using the Benjamini & Hochberg method and a false discovery rate of 10%, this P-value for interaction did not remain significant (critical threshold of significance for family of P-values was <0.03). (3) deseasonalised values.

**Table 4.** Mean VO_2peak_ in exercise sub-study participants at 1-, 2- and 3-year follow-up by allocation: overall and by sub-group.

|  |  |  | Vitamin D arm:<br>mean VO<sub>2peak</sub><br>ml/kg/min<br>(s.d.) [n] | Placebo arm:<br>mean VO<sub>2peak</sub><br>ml/kg/min<br>(s.d.) [n] | Adjusted mean<br>difference (95%<br>CI) <sup>(1)</sup> | P for<br>timepoint | Overall<br>P | P for<br>interaction |
| --- | --- | --- | --- | --- | --- | --- | --- | --- |
| Overall |  | 1<br>year | 43.2 (3.7)<br>[278] | 43.0 (3.4)<br>[296] | 0.27 (-0.30, 0.84) | 0.36 | 0.28 | -- |
|  |  | 2<br>years | 41.9 (3.9)<br>[268] | 41.9 (3.7)<br>[289] | 0.20 (-0.37, 0.78) | 0.48 |  |  |
|  |  | 3<br>years | 41.3 (4.2)<br>[268] | 41.3 (4.1)<br>[286] | 0.05 (-0.53, 0.62) | 0.87 |  |  |
| By sex | Male | 1<br>year | 45.0 (3.1)<br>[151] | 44.5 (3.1)<br>[144] | 0.61 (-0.09, 1.32) | 0.09 | 0.06 | 0.06 |
|  |  | 2<br>years | 43.9 (3.1)<br>[144] | 43.5 (3.3)<br>[143] | 0.55 (-0.17, 1.26) | 0.13 |  |  |
|  |  | 3<br>years | 43.6 (3.0)<br>[143] | 43.2 (3.8)<br>[143] | 0.46 (-0.25, 1.18) | 0.20 |  |  |
|  | Female | 1<br>year | 41.1 (3.3)<br>[127] | 41.6 (3.1)<br>[152] | -0.32 (-1.05, 0.40) | 0.38 | 0.47 |  |
|  |  | 2<br>years | 39.7 (3.5)<br>[124] | 40.2 (3.4)<br>[146] | -0.40 (-1.13, 0.32) | 0.28 |  |  |
|  |  | 3<br>years | 38.7 (3.8)<br>[125] | 39.4 (3.5)<br>[143] | -0.68 (-1.41, 0.04) | 0.07 |  |  |
| By baseline<br>25(OH)D<br>concentration <sup>(2)</sup> | <25<br>nmol/L | 1<br>year | 43.0 (4.2)<br>[103] | 43.0 (3.2)<br>[113] | -0.11 (-1.05, 0.83) | 0.82 | 0.44 | 0.06 |
|  |  | 2<br>years | 41.3 (4.0)<br>[104] | 41.9 (3.7)<br>[107] | -0.62 (-1.56, 0.32) | 0.20 |  |  |
|  |  | 3<br>years | 40.8 (4.6)<br>[100] | 41.3 (4.1)<br>[108] | -0.64 (-1.59, 0.30) | 0.18 |  |  |
|  | ≥25<br>nmol/L | 1<br>year | 43.4 (3.5)<br>[175] | 43.0 (3.5)<br>[183] | 0.50 (-0.22, 1.22) | 0.18 | 0.050 |  |
|  |  | 2<br>years | 42.4 (3.7)<br>[164] | 41.8 (3.7)<br>[182] | 0.71 (-0.02, 1.43) | 0.06 |  |  |
|  |  | 3<br>years | 41.6 (3.9)<br>[168] | 41.3 (4.1)<br>[178] | 0.46 (-0.26, 1.19) | 0.21 |  |  |
| By calcium<br>intake | <500<br>mg/day | 1<br>year | 43.2 (3.9)<br>[98] | 42.8 (3.5)<br>[102] | 0.36 (-0.41, 1.14) | 0.36 | 0.18 | 0.60 |
|  |  | 2<br>years | 42.0 (4.1)<br>[99] | 41.5 (3.7)<br>[102] | 0.48 (-0.30, 1.25) | 0.23 |  |  |
|  |  | 3<br>years | 41.2 (4.4)<br>[96] | 41.2 (4.2)<br>[102] | 0.13 (-0.64, 0.91) | 0.74 |  |  |
|  | ≥500<br>mg/day | 1<br>year | 43.5 (3.4)<br>[169] | 43.3 (3.5)<br>[174] | 0.27 (-0.68, 1.22) | 0.58 | 0.80 |  |
|  |  | 2<br>years | 42.1 (3.4)<br>[161] | 42.4 (3.7)<br>[179] | 0.01 (-0.94, 0.96) | 0.99 |  |  |
|  |  | 3<br>years | 41.5 (3.9)<br>[167] | 41.5 (4.0)<br>[176] | 0.08 (-0.88, 1.03) | 0.88 |  |  |

**Abbreviations:** VO<sub>2peak</sub>, peak oxygen consumption. 25(OH)D, 25-hydroxyvitamin D. CI, confidence interval. S.d., standard deviation. N, number.
**Footnotes** (1) adjusted for baseline value and school of attendance. (2) deseasonalised values.

**Table 5.**
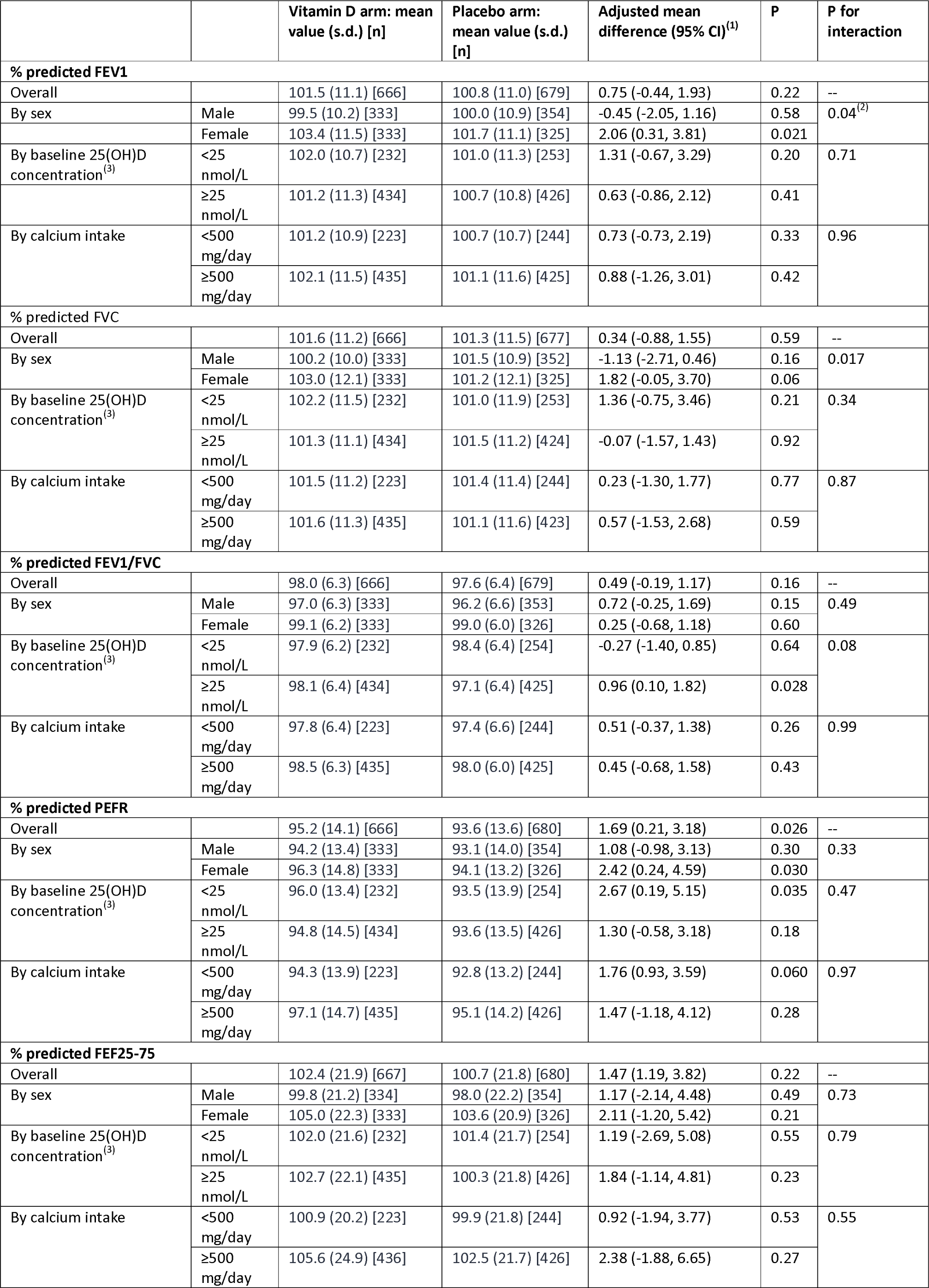
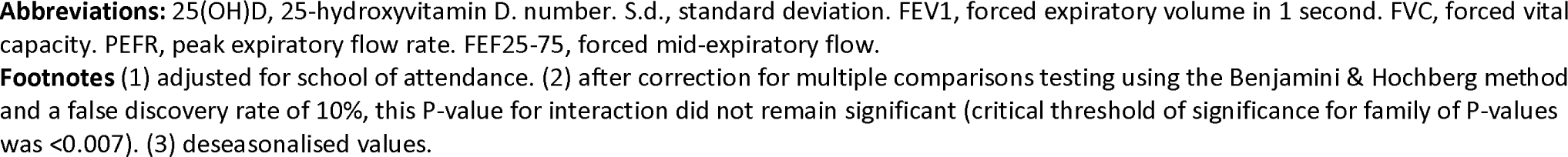
Spirometric outcomes at 3-year follow-up by allocation: overall and by sub-group.

|  |  | Vitamin D arm: mean value (s.d.) [n] | Placebo arm: mean value (s.d.) [n] | Adjusted mean difference (95% CI)<sup>(1)</sup> | P | P for interaction |
| --- | --- | --- | --- | --- | --- | --- |
| % predicted FEV1 |  |  |  |  |  |  |
| Overall |  | 101.5 (11.1) [666] | 100.8 (11.0) [679] | 0.75 (-0.44, 1.93) | 0.22 | -- |
| By sex | Male | 99.5 (10.2) [333] | 100.0 (10.9) [354] | -0.45 (-2.05, 1.16) | 0.58 | 0.04<sup>(2)</sup> |
|  | Female | 103.4 (11.5) [333] | 101.7 (11.1) [325] | 2.06 (0.31, 3.81) | 0.021 |  |
| By baseline 25(OH)D concentration<sup>(3)</sup> | <25 nmol/L | 102.0 (10.7) [232] | 101.0 (11.3) [253] | 1.31 (-0.67, 3.29) | 0.20 | 0.71 |
|  | ≥25 nmol/L | 101.2 (11.3) [434] | 100.7 (10.8) [426] | 0.63 (-0.86, 2.12) | 0.41 |  |
| By calcium intake | <500 mg/day | 101.2 (10.9) [223] | 100.7 (10.7) [244] | 0.73 (-0.73, 2.19) | 0.33 | 0.96 |
|  | ≥500 mg/day | 102.1 (11.5) [435] | 101.1 (11.6) [425] | 0.88 (-1.26, 3.01) | 0.42 |  |
| % predicted FVC |  |  |  |  |  |  |
| Overall |  | 101.6 (11.2) [666] | 101.3 (11.5) [677] | 0.34 (-0.88, 1.55) | 0.59 | -- |
| By sex | Male | 100.2 (10.0) [333] | 101.5 (10.9) [352] | -1.13 (-2.71, 0.46) | 0.16 | 0.017 |
|  | Female | 103.0 (12.1) [333] | 101.2 (12.1) [325] | 1.82 (-0.05, 3.70) | 0.06 |  |
| By baseline 25(OH)D concentration<sup>(3)</sup> | <25 nmol/L | 102.2 (11.5) [232] | 101.0 (11.9) [253] | 1.36 (-0.75, 3.46) | 0.21 | 0.34 |
|  | ≥25 nmol/L | 101.3 (11.1) [434] | 101.5 (11.2) [424] | -0.07 (-1.57, 1.43) | 0.92 |  |
| By calcium intake | <500 mg/day | 101.5 (11.2) [223] | 101.4 (11.4) [244] | 0.23 (-1.30, 1.77) | 0.77 | 0.87 |
|  | ≥500 mg/day | 101.6 (11.3) [435] | 101.1 (11.6) [423] | 0.57 (-1.53, 2.68) | 0.59 |  |
| % predicted FEV1/FVC |  |  |  |  |  |  |
| Overall |  | 98.0 (6.3) [666] | 97.6 (6.4) [679] | 0.49 (-0.19, 1.17) | 0.16 | -- |
| By sex | Male | 97.0 (6.3) [333] | 96.2 (6.6) [353] | 0.72 (-0.25, 1.69) | 0.15 | 0.49 |
|  | Female | 99.1 (6.2) [333] | 99.0 (6.0) [326] | 0.25 (-0.68, 1.18) | 0.60 |  |
| By baseline 25(OH)D concentration<sup>(3)</sup> | <25 nmol/L | 97.9 (6.2) [232] | 98.4 (6.4) [254] | -0.27 (-1.40, 0.85) | 0.64 | 0.08 |
|  | ≥25 nmol/L | 98.1 (6.4) [434] | 97.1 (6.4) [425] | 0.96 (0.10, 1.82) | 0.028 |  |
| By calcium intake | <500 mg/day | 97.8 (6.4) [223] | 97.4 (6.6) [244] | 0.51 (-0.37, 1.38) | 0.26 | 0.99 |
|  | ≥500 mg/day | 98.5 (6.3) [435] | 98.0 (6.0) [425] | 0.45 (-0.68, 1.58) | 0.43 |  |
| % predicted PEFR |  |  |  |  |  |  |
| Overall |  | 95.2 (14.1) [666] | 93.6 (13.6) [680] | 1.69 (0.21, 3.18) | 0.026 | -- |
| By sex | Male | 94.2 (13.4) [333] | 93.1 (14.0) [354] | 1.08 (-0.98, 3.13) | 0.30 | 0.33 |
|  | Female | 96.3 (14.8) [333] | 94.1 (13.2) [326] | 2.42 (0.24, 4.59) | 0.030 |  |
| By baseline 25(OH)D concentration<sup>(3)</sup> | <25 nmol/L | 96.0 (13.4) [232] | 93.5 (13.9) [254] | 2.67 (0.19, 5.15) | 0.035 | 0.47 |
|  | ≥25 nmol/L | 94.8 (14.5) [434] | 93.6 (13.5) [426] | 1.30 (-0.58, 3.18) | 0.18 |  |
| By calcium intake | <500 mg/day | 94.3 (13.9) [223] | 92.8 (13.2) [244] | 1.76 (0.93, 3.59) | 0.060 | 0.97 |
|  | ≥500 mg/day | 97.1 (14.7) [435] | 95.1 (14.2) [426] | 1.47 (-1.18, 4.12) | 0.28 |  |
| % predicted FEF25-75 |  |  |  |  |  |  |
| Overall |  | 102.4 (21.9) [667] | 100.7 (21.8) [680] | 1.47 (1.19, 3.82) | 0.22 | -- |
| By sex | Male | 99.8 (21.2) [334] | 98.0 (22.2) [354] | 1.17 (-2.14, 4.48) | 0.49 | 0.73 |
|  | Female | 105.0 (22.3) [333] | 103.6 (20.9) [326] | 2.11 (-1.20, 5.42) | 0.21 |  |
| By baseline 25(OH)D concentration<sup>(3)</sup> | <25 nmol/L | 102.0 (21.6) [232] | 101.4 (21.7) [254] | 1.19 (-2.69, 5.08) | 0.55 | 0.79 |
|  | ≥25 nmol/L | 102.7 (22.1) [435] | 100.3 (21.8) [426] | 1.84 (-1.14, 4.81) | 0.23 |  |
| By calcium intake | <500 mg/day | 100.9 (20.2) [223] | 99.9 (21.8) [244] | 0.92 (-1.94, 3.77) | 0.53 | 0.55 |
|  | ≥500 mg/day | 105.6 (24.9) [436] | 102.5 (21.7) [426] | 2.38 (-1.88, 6.65) | 0.27 |  |

**Abbreviations:** 25(OH)D, 25-hydroxyvitamin D. number. S.d., standard deviation. FEV1, forced expiratory volume in 1 second. FVC, forced vital capacity. PEFR, peak expiratory flow rate. FEF25-75, forced mid-expiratory flow.
**Footnotes** (1) adjusted for school of attendance. (2) after correction for multiple comparisons testing using the Benjamini & Hochberg method and a false discovery rate of 10%, this P-value for interaction did not remain significant (critical threshold of significance for family of P-values was <0.007). (3) deseasonalised values.

## DISCUSSION

We present findings of the largest randomised controlled trial to investigate effects of vitamin D on muscle strength, peak oxygen uptake and spirometric lung volumes in children. Vitamin D deficiency was highly prevalent among the study population at baseline, and the intervention was highly effective in elevating serum 25(OH)D concentrations among participants who were randomised to receive it. However, this was not associated with any effect on any physiological outcome investigated, either overall or in sub-groups defined by baseline vitamin D status, sex or calcium intake.

Our findings contrast with those of observational studies reporting associations between low vitamin D status and reduced muscle strength and cardiorespiratory fitness,^12^ ^13^ and chime with results of smaller RCTs, conducted in populations with lower prevalence of vitamin D deficiency, that have yielded null results.^18–22^ They are also consistent with the lack of effect seen for muscle strength and exercise outcomes in a similar trial conducted in South Africa,^35^ and for other ‘non-classical’ outcomes in trials of weekly vitamin D supplementation in children.^23–25^ ^36–38^ Inconsistency between positive findings of observational vs. null findings from interventional studies may reflect effects of confounding or bias in the former.^39^ Lack of sub-group effects in participants with lower baseline vitamin D status or calcium intake suggests that neither of these factors modified the effects of vitamin D supplementation on outcomes investigated. We highlight that our null findings do not have relevance for children with symptomatic vitamin D deficiency, since those who were found to have signs of rickets were excluded from the trial, as it would not have been ethical to randomise them to placebo.

Our study has several strengths. The large sample size and low rates of loss to follow-up maximised our power to detect effects of the intervention, and the high prevalence of vitamin D deficiency and low calcium intake at baseline allowed us to rule out effects even in sub-groups who might have been expected to derive particular benefit from vitamin D replacement. The intervention was highly effective in elevating serum 25(OH)D concentrations into the physiological range. We also assessed a comprehensive range of outcomes relating to muscle and cardiorespiratory fitness, with assessments at more than one time-point to allow detection of any effects that may have differed with varying duration of supplementation.

Our study also has some limitations. Spirometry was assessed at 3-year follow-up only: accordingly, analyses testing the effect of allocation to vitamin D vs. placebo could not be adjusted for baseline. However, we have no reason to suspect significance imbalance in baseline values between groups, as the randomisation process was effective in distributing all other baseline characteristics evenly between children randomised to intervention vs. control arms. We also highlight that our findings relate to effects of weekly vitamin D supplementation specifically. It remains technically possible that daily supplementation might have a different effect, although the fact that weekly supplementation is effective in suppressing PTH and ALP concentrations in study participants^25^ ^37^ suggests that this dosing regimen exerts the same physiological effects as would be expected with daily supplementation.

In conclusion, this large multicentre RCT of vitamin D supplementation, administered for 3 years to a population of children with low baseline 25(OH)D concentrations, did not show any effect of the intervention on muscle strength, cardiorespiratory fitness or spirometric outcomes. Taken together with null results from a similarly-designed Phase 3 RCT conducted in CapeTown, South Africa,^35^ our study does not suggest a role for weekly oral vitamin D supplementation to enhance muscle strength, peak oxygen uptake or respiratory function in schoolchildren in whom rickets has been excluded.

## Acknowledgements

We thank all the children who participated in the trial, and their parents and guardians. We also thank independent members of the Data Safety Monitoring Board (Prof S.M. Fortune and Dr P.L. Williams, Harvard T.H. Chan School of Public Health; Profs M.F. Holick and C.R. Horsburgh, Boston University; Prof P. Enkhbaatar, University of Texas; and Dr. E. Chadraa, Minnesota State University); independent members of the Trial Steering Committee (Profs W.C. Willett, Edward L. Giovannucci and B.R. Bloom, Harvard T.H. Chan School of Public Health; Dr N. Naranbat, Gyals Medical Laboratory, Ulaanbaatar; and late Dr D. Malchinkhuu, National Center for Maternal and Child Health of Mongolia); board members at the Mongolian Health Initiative (Dr J. Tuyatsetseg, Mongolian University of Science and Technology; Dr J. Amarsanaa, Happy Veritas Laboratory, Ulaanbaatar; Drs P. Erkhembulgan and G. Batbaatar, Mongolian National University of Medical Sciences; Prof M.C. Elliott, Harvard University; and Mr. Ts. Munh-Orgil, Member of the Mongolian Parliament); and the following individuals for advice and helpful discussions: Dr Winthrop Burr (Anadyne Psychotherapy Inc) and Dr. Masae Kawamura at Qiagen USA.

## Contributors Contributors

DG and ARM conceived the study and contributed to study design and protocol development; SH contributed to design of outcome assessments of muscle strength and peak oxygen uptake. DG led on trial implementation, with support from UB, GG, UA, TT, KD, ET, SE, NE, C-E A, YT, TB, EG and EP. ET performed and supervised measurements of serum 25(OH)D concentrations. DG and ARM drafted the statistical analysis plan. DG, UB, ET and SE managed data. DG, ARM and DAJ accessed and verified the data underlying the study. DAJ conducted statistical analyses. ARM, DG and DAJ wrote the first draft of the trial report. All other authors made substantive comments thereon and approved the final version for submission.

## Patient consent for publication

Not applicable.

## Ethics approval

The study was approved by institutional review boards of the Mongolian Ministry of Health, Mongolian National University, and Harvard T. H. Chan School of Public Health (IRB # 14-0513). Participants and their parents/guardians provided written informed assent and consent, respectively, to take part in the trial before any study procedures were conducted.

## Data availability statement

Anonymised data are available from corresponding authors upon reasonable request, subject to terms of IRB and regulatory approval.

